# Lymphovascular Invasion Detection in Breast Cancer Using Deep Learning

**DOI:** 10.1101/2025.06.07.25329195

**Authors:** Hassan Keshvarikhojasteh, Nikolas Stathonikos, Paul Pham, Paul J. van Diest, Celien Vreuls, Christof A. Bertram, Josien P.W. Pluim, Mitko Veta

## Abstract

Lymphovascular invasion (LVI) is a critical pathological feature in breast cancer, strongly associated with an increased risk of metastasis and poorer prognosis. However, manual detection of LVI is labor-intensive and prone to inter-observer variability. To address these challenges, this study explores the potential of Swin-Transformer, a state-of-the-art deep learning model, and GigaPath, a cutting-edge foundation model, for automating the detection of LVI in whole-slide images (WSIs) of breast cancer tissue. We trained the models on a dataset of 90 annotated Hematoxylin and Eosin (H&E)-stained breast cancer slides, achieving strong performance with a slide-level Area Under the Receiver Operating Characteristic (AUC) of 97%, a sensitivity of 79% with an average of 8 false positives (FPs) per slide using the best-performing model. The results underscore the potential of Swin-Transformer and GigaPath to enhance diagnostic accuracy and consistency in LVI detection.

## Introduction

Breast cancer is the most commonly diagnosed cancer and the leading cause of cancer-related mortality among women worldwide. In 2020, it was estimated that 2.3 million new cases of breast cancer were diagnosed globally, surpassing lung cancer and accounting for 11.7% of all new cancer cases, making it a significant public health concern [1]. Early detection and accurate diagnosis are essential for improving patient outcomes, as they enable timely diagnosis and effective treatment interventions [2]. Breast cancer is a highly heterogeneous disease between and within tumors, encompassing various subtypes with distinct molecular and clinical characteristics, which further complicates its diagnosis and management [3]. This heterogeneity necessitates precise and reliable diagnostic tools to guide personalized treatment strategies, ultimately improving prognosis and survival rates [4].

Among the critical pathological features of breast cancer is lymphovascular invasion (LVI), which is defined as the presence of cancer cells within the lymphatic system and/or blood vessels [5]. The detection of LVI is strongly associated with an increased risk of metastasis and a poorer prognosis, making it an important factor in disease staging and therapeutic decision-making systems [6]. Traditionally, LVI is detected through histopathological examination of tissue samples, typically stained with hematoxylin and eosin (H&E) or immunohistochemistry (IHC) markers [7]. However, the visual identification of LVI by pathologists on H&E-stained slides is a complex and time-consuming process prone to inter-observer variability, leading to inconsistencies in diagnosis and treatment planning [8]. This variability underscores the need for more objective, reliable, and automated methods for detecting LVI in breast cancer.

Digital pathology has revolutionized the field through the digitization of histopathological slides, facilitating storage, analysis, education, and the sharing of pathology data in a digital format. At its core is digitizing microscopic entire slides into high-resolution digital whole-slide images (WSI) [9]. WSI facilitate more efficient workflows in clinical, research, and educational settings by enabling pathologists to review slides remotely, perform image analysis using advanced computational tools, and easily share data for second opinions or collaborative studies [10]. Moreover, the adoption of WSI has paved the way for the integration of artificial intelligence (AI) in pathology, allowing the development of AI-powered diagnostic tools that can assist the pathologist in tasks such as tumor detection, grading, and quantification of pathological features [11–13].

AI is a broad field that aims to replicate human intelligence in computational systems. One of the most powerful approaches within AI is deep learning, a technique that enables models to automatically learn complex patterns from large-scale datasets. Deep learning has proven particularly effective for handling high-dimensional data, and its success has led to significant advancements across a range of domains, providing the foundation for many AI-driven analysis and decision support systems.

In recent years, deep learning, particularly convolutional neural networks (CNNs), has transformed medical image analysis, offering new possibilities for automating diagnostic tasks [14]. CNNs have demonstrated exceptional performance in a wide range of medical imaging tasks, including tumor detection, segmentation, and classification [15]. Their ability to automatically learn hierarchical features from raw image data has made them a powerful tool for analyzing complex medical images.

However, CNNs have limitations, particularly when applied to complex histopathological images. Specifically, CNNs often struggle to capture spatial long-range dependencies—such as relationships between distant regions within a large tissue sample—as well as hierarchical structures, where patterns at different scales (e.g., cellular, tissue, and organ levels) are important for diagnosis. For example, in pathology, the interaction between tumor cells and lymphatic vessels, which may occur across distant regions, and the organization of cells into higher-order tissue structures both provide critical diagnostic information that can be difficult for standard CNNs to model effectively [16]. To address these challenges, researchers have increasingly explored Transformer-based models like Vision Transformer (ViT) which have shown great promise in computer vision applications [17]. ViTs leverage self-attention mechanisms, which enable the model to compare and relate different parts of an input image to each other. Specifically, self-attention computes the relevance of each image patch with respect to all other patches, allowing the model to capture global context and long-range dependencies. This capability makes ViTs particularly effective for tasks that require understanding complex spatial relationships within high-resolution images.

However, while ViTs have demonstrated impressive capabilities, they often require extensive computational resources and large-scale datasets, which can limit their applicability in certain medical imaging contexts. Building on the strengths of ViTs while addressing their limitations, the Swin-Transformer (Shifted Window Transformer) was introduced as a more data-efficient and scalable variant of the Transformer architecture specifically designed for vision tasks [18].

Foundation models (FMs) are large-scale models trained on large-scale and diverse datasets, designed to learn general-purpose representations that can be adapted to a wide range of downstream tasks. Unlike traditional models that are usually trained for a specific task, FMs aim to provide a strong starting point (“foundation”) for solving various problems with minimal additional training. These models often leverage self-supervised learning (SSL) techniques, where the model learns patterns and structures in data without requiring manual labels. For example, a model might be trained to predict missing parts of an image. This approach enables FMs to capture rich and transferable features from histological tissues, making them particularly valuable for clinical applications where labeled data is scarce [19, 20]. The success of FMs is largely attributed to Transformer-based architectures, which effectively integrate global context [21]. GigaPath [22] is a newly developed foundation model for whole-slide pathology, pretrained on a diverse collection of whole-slide images from a large U.S. healthcare network. GigaPath uses a novel vision transformer architecture and integrates the LongNet method [23] to efficiently process slide-level representations across tens of thousands of image tiles. As an open-weight foundation model, GigaPath achieves state-of-the-art performance across multiple digital pathology tasks, demonstrating its potential for advancing computational pathology.

Recently, two methods have been successfully developed for detecting LVI in testicular [24] and gastric cancers [25]. These approaches utilize patch-level object detection and classification for LVI identification at the slide level. However, accurate annotation of LVI foci is crucial for object detection.

In this study, we train a Swin-Transformer (Swin-Small) model as a patch-level classifier on breast cancer slides. Additionally, we explore the use of a foundation model, GigaPath, to detect the location of LVI, an area that has not been extensively researched.

## Materials and methods

### Ethics statement

This study involved the retrospective analysis of fully anonymized H&E-stained breast cancer slides obtained from the University Medical Center Utrecht (UMCU). According to the Medical Ethics Review Committee (METC) of UMC Utrecht, the use of anonymized, retrospective data does not require ethical approval or informed consent. No identifiable information was accessed by the authors during or after data collection. The slides were accessed for research purposes in April 2024. All experiments were carried out in accordance with the Declaration of Helsinki and the International Ethical Guidelines for Biomedical Research Involving Human Subjects by the Council for International Organizations of Medical Sciences (CIOMS) [26, 27].

### Data

We retrieved 90 H&E-stained breast cancer slides from UMCU. These slides were scanned for routine diagnostics, each containing at least one focus of LVI, from a cohort of 67 patients (median: 1.3 slides per patient, range: 1–3). The slides were scanned using a Hamamatsu XR nanozoomer 2.0 scanner (Hamamatsu Photonics, Japan) at a magnification of 40 *×*, corresponding to a resolution of 0.2263 *µm/pixel*. Comprehensive pathological annotation was performed, including histological subtype and molecular profiling (Table 1). Invasive carcinoma of no special type (NST) was the predominant histological subtype in the cohort, accounting for 77.6% (n = 52). Molecular subtyping showed a predominance of Luminal A (ER+/PR+/HER2-) tumors (79.1%, n=53), followed by triple-negative (ER-/PR-/HER2-) tumors (16.4%, n=11).

**Table 1.** Clinicopathological characteristics of the breast cancer cohort (n = 67) evaluated for lymphovascular invasion.

| Characteristic | n (%) | Notes |
| --- | --- | --- |
| *Histological Subtype* |  | WHO classification |
| Invasive carcinoma NST* | 52 (77.6%) |  |
| Infiltrating ductulolobular | 10 (14.9%) |  |
| Other | 5 (7.5%) | • Ductulolobular (n=2)<br>• Metaplastic (n=1)<br>• Invasive lobular (n=1)<br>• Invasive ductulolobular (n=1) |
| *Molecular Subtype* |  | IHC/FISH-defined |
| Luminal A (ER+/PR+/HER2<sup>-</sup>) | 53 (79.1%) |  |
| Triple-negative | 11 (16.4%) |  |
| Other | 3 (4.5%) | • HER2-enriched (n=1)<br>• Luminal B (n=2) |

\* NST = No Special Type.

LVI was recognized according to standard histological criteria: usually rounded smaller groups of tumor cells located in a vascular space lined with endothelium, while allowing space between tumor cells and endothelium, usually located outside the dominant tumor nodule or between benign breast structures, often in the vicinity of other vascular structures or nerves. Two board-certified pathologists, who were blinded to each other’s annotations, independently provided bounding box annotations of variable size, each approximately encompassing the LVI foci on the slides. In cases of disagreement, a third board-certified pathologist reviewed the annotations and made the final determination. This process resulted in a total of 660 annotated LVI foci. The dataset was divided into training, validation, and testing sets using a 6:2:2 ratio, ensuring that slides from the same patient were not assigned to different sets. An overview of the framework is shown in Fig 1.

**Fig 1.**
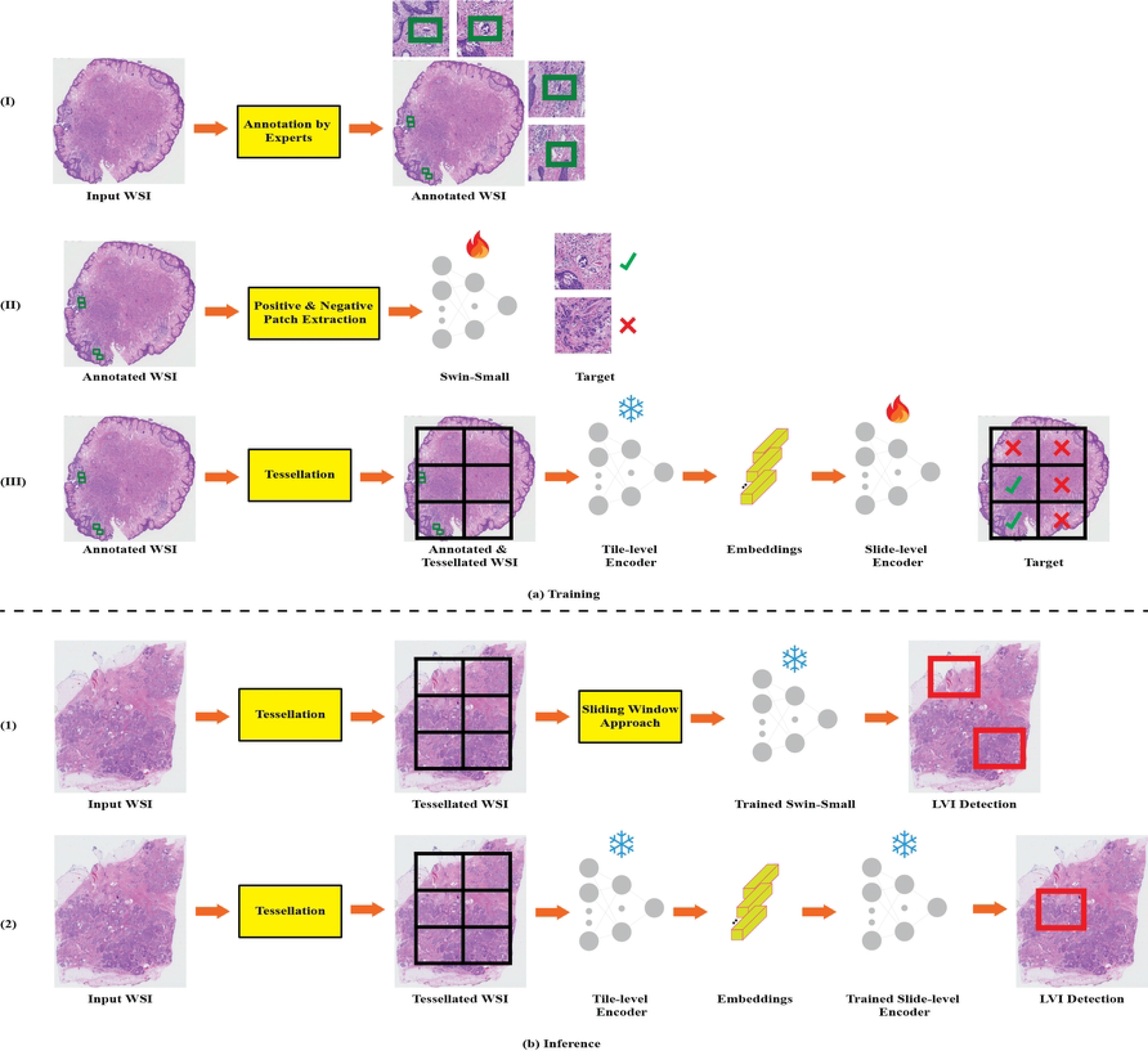
Illustration of the Proposed Framework (a) Training Phase: (I) Lymphovascular invasion (LVI) foci in breast cancer are identified through consensus annotations by two expert pathologists, with a third pathologist resolving any disagreements. (II) For the Swin-Small model, image patches containing LVI-positive and LVI-negative regions are extracted based on these annotations and used for model fine-tuning. (III) In contrast, for GigaPath, the whole slide image (WSI) is tessellated, and patch embeddings are extracted using a frozen tile-level encoder. The slide-level encoder is subsequently fine-tuned for binary classification of all patches. (b) Inference Phase: (1) For Swin-Small, the input WSI undergoes tessellation, followed by a sliding window approach to predict patch-wise probabilities. Postprocessing techniques (described in Section) are then applied to highlight patches with high probabilities as potential LVI locations. (2) For GigaPath, after tessellation, patch features are extracted using the tile-level encoder, and probabilities for all patches are computed in a single step using the slide-level encoder. LVI locations are subsequently identified using the same postprocessing techniques.

For the Swin-Small model, the slides were tessellated into tiles of 512 *×* 512 pixels at 10*×* magnification (0.9052 *µm/pixel*), with a 50% overlap between adjacent patches. The tile size was selected based on the average LVI rectangle sizes observed in the training set. LVI-positive patches were identified based on the ground-truth annotations, while non-LVI patches were randomly sampled from the remaining areas. Specifically, for each annotated LVI foci, we extracted a fixed number of positive patches (n=40) centered around the annotation. Negative patches were extracted from regions not containing LVI, with the number of negative patches constituting 25% of the total patches on a slide. These configurations were designed to ensure a sufficient collection of positive and negative patches while accounting for the class imbalance. To ensure compatibility with the model input size, all patches were resized to 224 *×* 224 pixels.

For GigaPath, we first extracted the embeddings of the patches at level 1 (20*×* magnification, 0.4526 *µm/pixel*, 1024 *×* 1024 pixels) as recommended by the developers using the tile-level encoder. Subsequently, we fine-tuned the slide-level encoder. During this process, after tessellation, tiles containing LVI foci were considered positive, while the remaining tiles were labeled as negative.

### Training

We trained a Swin-Small network for the binary classification of patches, distinguishing between LVI and non-LVI ones. Starting with an ImageNet pre-trained model, we replaced the final classification layer to suit our binary task and fine-tuned all layers of the network. To address the issue of class imbalance, we used a weighted loss function instead of random undersampling, as it yielded better performance. The model was trained for 15 epochs including one warmup epoch with a batch-size of 64. We used AdamW optimizer, incorporating cosine scheduler, with an initial learning_rate of 1e-1. Moreover, we set weight_decay= 2e-3 as a regularization technique. We selected the best model according to the F1 score. Based on the initial experiment, where no data augmentation techniques were used, we found that applying a horizontal flip with a probability of 0.5 was beneficial. In addition, color_jitter= 0.4 and using the default values for scale and ratio further enhanced the model’s performance. An example of one input batch is displayed in Fig 2.

**Fig 2.**
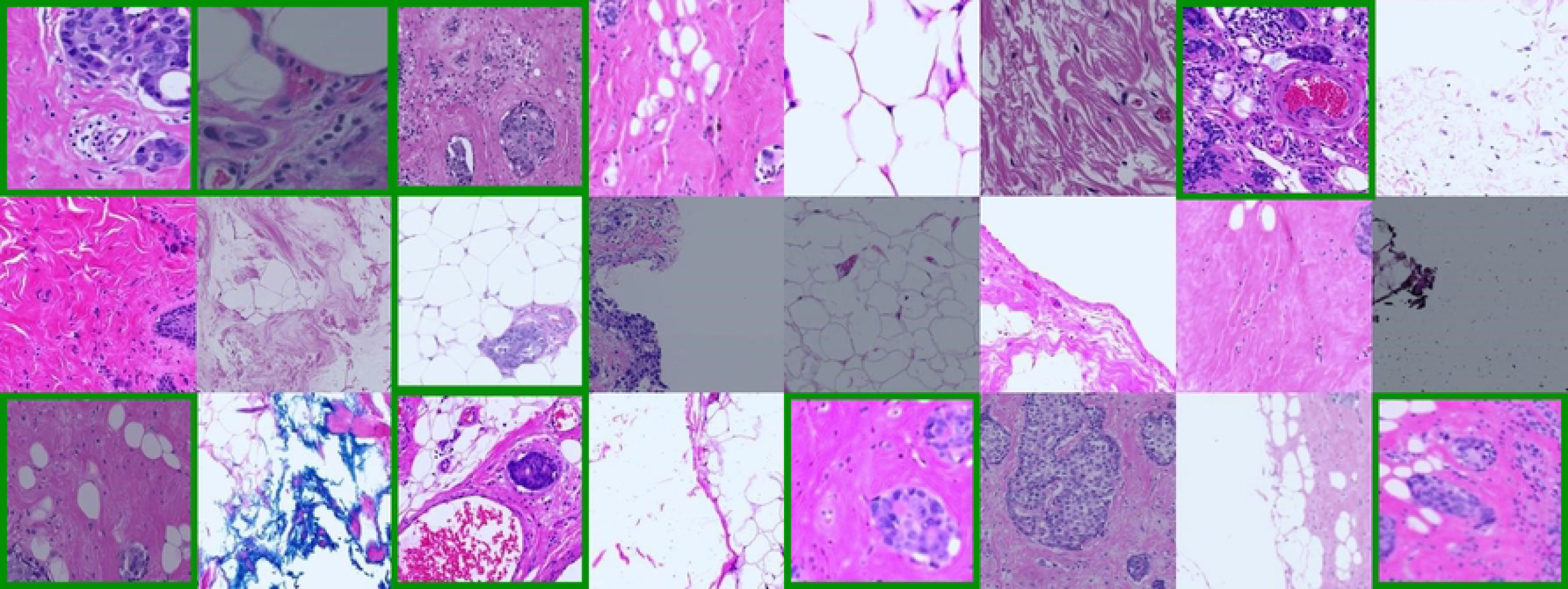
A randomly selected input batch fed to the Swin-Small model, with applied data augmentation techniques. The lymphovascular invasion-positive patches are outlined with green rectangles.

For training the slide-level encoder of GigaPath, we used a weighted loss function that accounts for the proportion of LVI-positive and LVI-negative patches within each WSI. The slide-level encoder was fine-tuned for 15 epochs using a learning_rate of 1e-3 and a weight_decay of 1e-2. We monitored the Area Under the Free-Response Receiver Operating Characteristic (FROC) curve for maximum of 22 average False Positives (FPs) to select the optimal model, as sensitivity beyond this threshold is considered unimportant. To improve generalization, we applied slide-level data augmentation rather than patch-level augmentation. Specifically, we first determined random color jitter parameters for the input WSI and then applied them consistently across all extracted patches. Fig 3 illustrates a WSI alongside its augmented counterparts.

**Fig 3.**
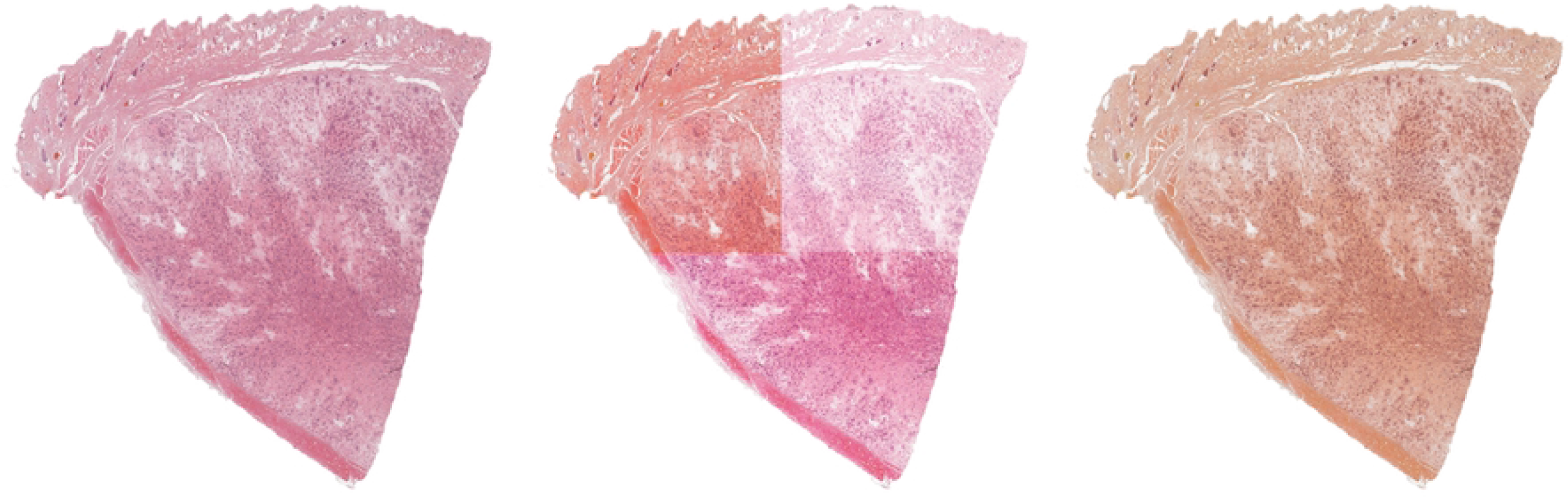
From left to right: A whole-slide image (WSI), the WSI after applying patch-level data augmentation, and the WSI after applying slide-level data augmentation. This illustration considers only three patches after tessellation for clarity and convenience.)

### Inference

During inference, we used a sliding-window approach for the Swin-Small network in which we predicted the LVI probability for each tile in the input WSI using the best-selected model. However, for GigaPath, we generated the LVI probabilities of whole patches within a WSI in a single-step using the slide-level encoder. We then highlighted the patches with probabilities exceeding the optimal threshold, defined on the validation set. Given that the tiles were extracted with 50% overlap, we merged the overlapping high-probability patches to refine the final detection.

### Statistical analysis

We evaluated the models’ performance at slide level. We reported the average Area Under the Receiver Operating Characteristic (AUC), True Positive Rate (TPR) and number of FPs. Moreover, we display the FROC curve for maximum of 22 average FPs.

## Results

### Slide-wise

As detailed in *Inference* section, all tiles of the input WSI were classified according to the optimal thresholds. The slide-level results are summarized in Table 2. In addition, we also display FROC curve for the test set in Fig 4. The results demonstrated that GigaPath outperformed Swin-Small.

**Table 2.** The slide-level results of two models for detecting lymphovascular invasion in breast cancer.

| Model | AUC (↑) | TPR (↑) | FP (↓) |
| --- | --- | --- | --- |
| Swin-Small | 0.95 | 0.75 | 14 |
| **GigaPath** | **0.97** | **0.79** | **8** |

**Fig 4.**
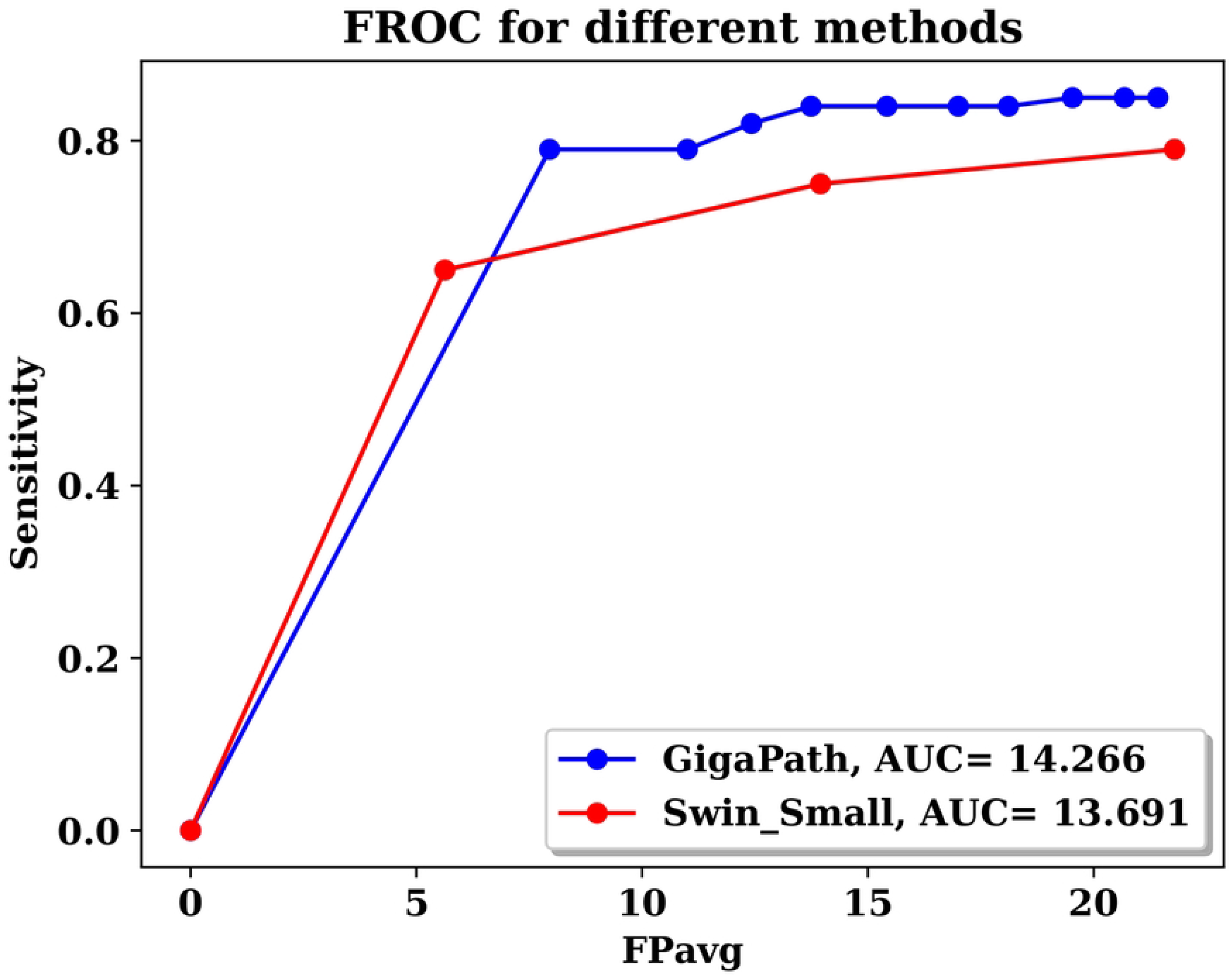
FROC curves for Swin-Small and GigaPath lymphovascular invasion detection models for maximum of 22 average false positives.

### False negatives and positives

For qualitative analysis, the predictions of both models for two representative WSIs are visualized in Fig 5 and Fig 6. In Fig 5, the Swin-Small model demonstrated 50% sensitivity in detecting LVI, correctly identifying one of two LVI foci but generating several FP predictions. In contrast, GigaPath outperformed Swin-Small, accurately detecting both LVI foci with zero FPs. Moreover in Fig 6, when challenged with subtle or weakly expressed LVI foci, the limitations of Swin-Small become more apparent, as it fails to detect any of the four LVI foci while still producing false-positive artifacts. GigaPath, though not flawless, shows greater robustness by correctly identifying two of the four LVI foci. Its performance trade-off includes a higher FP rate compared to Swin-Small in this specific example, suggesting room for optimization in low-signal contexts.

**Fig 5.**
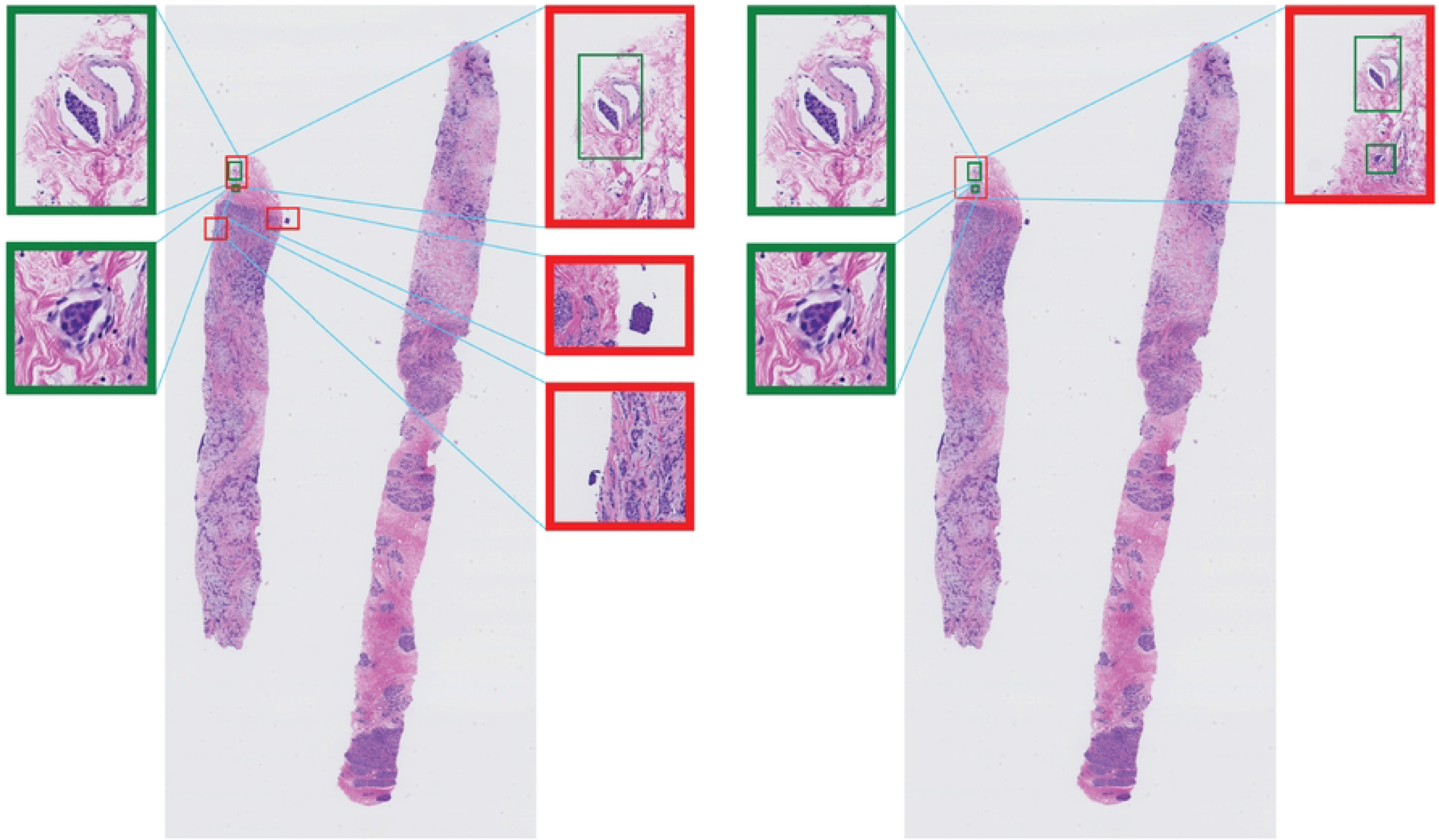
Left: A whole slide image (WSI) with predictions from Swin-Small. Right: The same WSI with predictions from GigaPath. The predicted lymphovascular invasion (LVI) locations are highlighted with red boxes, while the ground-truth LVI locations are marked with green boxes.

**Fig 6.**
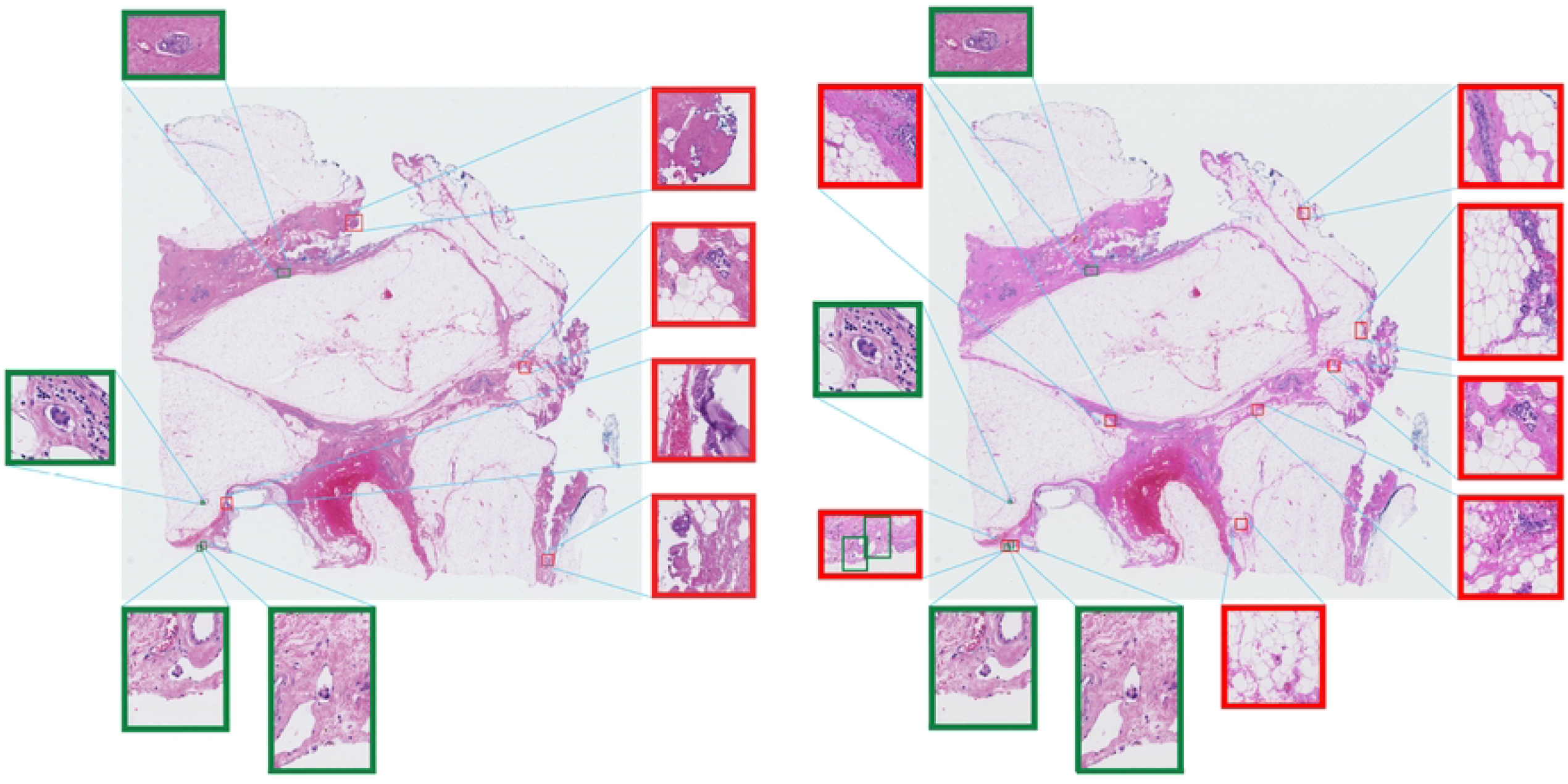
Left: A WSI with predictions from Swin-Small. Right: The same WSI with predictions from GigaPath. The predicted LVI locations are highlighted with red boxes, while the ground-truth LVI locations are marked with green boxes.

## Discussion & Conclusion

In this study, we evaluated the effectiveness of transformer-based and foundation models for LVI detection in breast cancer WSIs. Our results indicate that GigaPath, a foundation model trained on a diverse histopathology dataset, outperforms Swin-Small at the slide level, demonstrating superior generalization. Swin-Small’s overall slide-level classification remained lower, indicating that foundation models benefit from broader domain exposure and large-scale weak supervision. This finding underscores the importance of both model architecture and dataset diversity in achieving high-performance in histopathology.

One key challenge observed in our analysis was the occurrence of false positives, particularly in areas with staining artifacts or structures mimicking LVI such as retraction artefacts, tissue folds or debris. This finding underscores the importance of combining automated tools with expert review, as pathologists can distinguish between true LVIs and easily identifiable artifacts that the model might mistakenly identify as positive. However, such artefacts are usually recognized by pathologists, and even an algorithm with a limited number of FPs may save the pathologist significant time in detecting LVI, especially because only one true positive LVI foci is required to establish the diagnosis; the number of LVI foci is not considered [6]. Addressing this issue could involve refining the model further or incorporating additional preprocessing steps to reduce the influence of such artifacts. Our findings align with prior studies that emphasize the effectiveness of self-supervised and transformer-based models in digital pathology [28].

From a clinical perspective, improving automated LVI detection could have significant implications for patient prognosis and treatment planning. GigaPath’s stronger performance at the slide level suggests that foundation models trained on large, diverse datasets could serve as valuable tools in pathology workflows. However, further validation across multi-center datasets is needed to assess their reliability in real-world clinical settings.

Future work should focus on expanding the dataset to include a more diverse range of patient samples, including those from multiple scanners or centers, as well as different histological subtypes of breast cancer [29]. This may not only improve the generalizability of the model but also allow for the exploration of its performance across different cancer subtypes, which may present varying patterns of LVI. Additionally, integrating multimodal data, such as combining WSI with molecular or genomic data, could further enhance the accuracy and clinical utility of the model. Another promising direction is the development of more efficient foundation models that require less computational power while maintaining high performance, making them more accessible for clinical settings.

In conclusion, this study demonstrates the efficacy of the transformer-based and foundation models in detecting LVI in breast cancer WSIs, offering a promising tool for improving the accuracy and consistency of LVI detection by pathologists. While further validation and refinement are necessary, the approach holds potential for significant benefits in clinical practice, particularly in standardizing and expediting the detection of LVI.

## Data Availability Statement

The dataset used in this study was obtained from UMCU and is not publicly available due to patient privacy regulations. It is available from the corresponding author upon reasonable request and with appropriate institutional approvals. All experiments were conducted using PyTorch on a high-performance computing system provided by UMCU. The source code for the framework is publicly available at https://github.com/tueimage/LVI-Detection.

## Acknowledgments

This work was done as a part of the IMI BigPicture project (IMI945358).

## Author Contributions

Conceptualization: Hassan Keshvarikhojasteh, Mitko Veta

Data curation: Hassan Keshvarikhojasteh, Paul J. van Diest, Celien Vreuls, Christof A. Bertram, Nikolas Stathonikos, Paul Pham

Formal analysis: Hassan Keshvarikhojasteh

Methodology: Hassan Keshvarikhojasteh, Mitko Veta

Supervision: Mitko Veta, Josien P.W. Pluim Writing – original draft: Hassan Keshvarikhojasteh

Writing – review & editing: Hassan Keshvarikhojasteh, Paul J. van Diest, Christof A.

Bertram, Nikolas Stathonikos, Paul Pham, Josien P.W. Pluim, Mitko Veta

## References

1. Sung H, Ferlay J, Siegel RL, Laversanne M, Soerjomataram I, Jemal A, et al. Global cancer statistics 2020: GLOBOCAN estimates of incidence and mortality worldwide for 36 cancers in 185 countries. CA: a cancer journal for clinicians. 2021;71(3):209–249.

2. Smith RA, Andrews KS, Brooks D, Fedewa SA, Manassaram-Baptiste D, Saslow D, et al. Cancer screening in the United States, 2019: A review of current American Cancer Society guidelines and current issues in cancer screening. CA: a cancer journal for clinicians. 2019;69(3):184–210.

3. Polyak K. Heterogeneity in breast cancer. The Journal of clinical investigation. 2011;121(10):3786–3788.

4. Simpson PT, Reis-Filho JS, Gale T, Lakhani SR. Molecular evolution of breast cancer. The Journal of Pathology: A Journal of the Pathological Society of Great Britain and Ireland. 2005;205(2):248–254.

5. Weigelt B, Geyer FC, Reis-Filho JS. Histological types of breast cancer: how special are they? Molecular oncology. 2010;4(3):192–208.

6. Rakha EA, Martin S, Lee AH, Morgan D, Pharoah PD, Hodi Z, et al. The prognostic significance of lymphovascular invasion in invasive breast carcinoma. Cancer. 2012;118(15):3670–3680.

7. Rakha EA, Reis-Filho JS, Baehner F, Dabbs DJ, Decker T, Eusebi V, et al. Breast cancer prognostic classification in the molecular era: the role of histological grade. Breast cancer research. 2010;12:1–12.

8. Hoda SA, Hoda RS, Merlin S, Shamonki J, Rivera M. Issues relating to lymphovascular invasion in breast carcinoma. Advances in anatomic pathology. 2006;13(6):308–315.

9. Pantanowitz L, Valenstein PN, Evans AJ, Kaplan KJ, Pfeifer JD, Wilbur DC, et al. Review of the current state of whole slide imaging in pathology. Journal of pathology informatics. 2011;2(1):36.

10. Evans AJ, Brown RW, Bui MM, Chlipala EA, Lacchetti C, Milner Jr DA, et al. Validating whole slide imaging systems for diagnostic purposes in pathology: guideline update from the College of American Pathologists in collaboration with the American Society for Clinical Pathology and the Association for Pathology Informatics. Archives of pathology & laboratory medicine. 2022;146(4):440–450.

11. Komura D, Ishikawa S. Machine learning methods for histopathological image analysis. Computational and structural biotechnology journal. 2018;16:34–42.

12. Aeffner F, Zarella MD, Buchbinder N, Bui MM, Goodman MR, Hartman DJ, et al. Introduction to digital image analysis in whole-slide imaging: a white paper from the digital pathology association. Journal of pathology informatics. 2019;10(1):9.

13. Hanna MG, Reuter VE, Hameed MR, Tan LK, Chiang S, Sigel C, et al. Whole slide imaging equivalency and efficiency study: experience at a large academic center. Modern Pathology. 2019;32(7):916–928.

14. Litjens G, Kooi T, Bejnordi BE, Setio AAA, Ciompi F, Ghafoorian M, et al. A survey on deep learning in medical image analysis. Medical image analysis. 2017;42:60–88.

15. Esteva A, Kuprel B, Novoa RA, Ko J, Swetter SM, Blau HM, et al. Dermatologist-level classification of skin cancer with deep neural networks. nature. 2017;542(7639):115–118.

16. Atabansi CC, Nie J, Liu H, Song Q, Yan L, Zhou X. A survey of Transformer applications for histopathological image analysis: New developments and future directions. BioMedical Engineering OnLine. 2023;22(1):96.

17. Dosovitskiy A, Beyer L, Kolesnikov A, Weissenborn D, Zhai X, Unterthiner T, et al. An image is worth 16×16 words: Transformers for image recognition at scale. arXiv preprint arXiv:201011929. 2020;.

18. Liu Z, Lin Y, Cao Y, Hu H, Wei Y, Zhang Z, et al. Swin transformer: Hierarchical vision transformer using shifted windows. In: Proceedings of the IEEE/CVF international conference on computer vision; 2021. p. 10012–10022.

19. Moor M, Banerjee O, Abad ZSH, Krumholz HM, Leskovec J, Topol EJ, et al. Foundation models for generalist medical artificial intelligence. Nature. 2023;616(7956):259–265.

20. Waqas A, Bui MM, Glassy EF, El Naqa I, Borkowski P, Borkowski AA, et al. Revolutionizing digital pathology with the power of generative artificial intelligence and foundation models. Laboratory investigation. 2023;103(11):100255.

21. Zhou C, Li Q, Li C, Yu J, Liu Y, Wang G, et al. A comprehensive survey on pretrained foundation models: A history from bert to chatgpt. International Journal of Machine Learning and Cybernetics. 2024; p. 1–65.

22. Xu H, Usuyama N, Bagga J, Zhang S, Rao R, Naumann T, et al. A whole-slide foundation model for digital pathology from real-world data. Nature. 2024;630(8015):181–188.

23. Ding J, Ma S, Dong L, Zhang X, Huang S, Wang W, et al. Longnet: Scaling transformers to 1,000,000,000 tokens. arXiv preprint arXiv:230702486. 2023;.

24. Ghosh A, Sirinukunwattana K, Khalid Alham N, Browning L, Colling R, Protheroe A, et al. The potential of artificial intelligence to detect lymphovascular invasion in testicular cancer. Cancers. 2021;13(6):1325.

25. Lee J, Cha S, Kim J, Kim JJ, Kim N, Jae Gal SG, et al. Ensemble deep learning model to predict lymphovascular invasion in gastric cancer. Cancers. 2024;16(2):430.

26. Collins GS, Reitsma JB, Altman DG, Moons KG. Transparent reporting of a multivariable prediction model for individual prognosis or diagnosis (TRIPOD) the TRIPOD statement. Circulation. 2015;131(2):211–219.

27. McShane LM, Altman DG, Sauerbrei W, Taube SE, Gion M, Clark GM. REporting recommendations for tumor MARKer prognostic studies (REMARK). Breast cancer research and treatment. 2006;100:229–235.

28. Chen RJ, Chen C, Li Y, Chen TY, Trister AD, Krishnan RG, et al. Scaling vision transformers to gigapixel images via hierarchical self-supervised learning. In: Proceedings of the IEEE/CVF conference on computer vision and pattern recognition; 2022. p. 16144–16155.

29. Aubreville M, Stathonikos N, Donovan TA, Klopfleisch R, Ammeling J, Ganz J, et al. Domain generalization across tumor types, laboratories, and species—insights from the 2022 edition of the mitosis domain generalization challenge. Medical Image Analysis. 2024;94:103155.

